# Increased rate of joint hypermobility in autism and related neurodevelopmental conditions is linked to dysautonomia and pain

**DOI:** 10.1101/2020.09.14.20194118

**Authors:** Jenny L L Csecs, Valeria lodice, Charlotte L Rae, Alice Brooke, Rebecca Simmons, Nicholas G Dowell, Fenella Prowse, Kristy Themelis, Hugo D Critchley, Jessica A Eccles

## Abstract

**Objective:** Autism, attention deficit hyperactivity disorder (ADHD), and tic disorder (Tourette syndrome; TS) are neurodevelopmental conditions that frequently co-occur and impact psychological, social and emotional functioning. Vulnerability to chronic physical symptoms, including fatigue and pain, are also recognised. The expression of joint hypermobility, reflecting a constitutional variant in connective tissue, predicts vulnerability to psychological symptoms alongside recognised physical symptoms. Here, we tested for increased rates of joint hypermobility, autonomic dysfunction and pain in 109 adults with neurodevelopmental diagnoses.

**Method:** Rates of generalized joint laxity in those individuals with neurodevelopmental conditions were compared to those in the general population in UK. Levels of orthostatic intolerance and musculoskeletal symptoms were compared to a neurotypical control group.

**Results:** Adults with neurodevelopmental diagnoses manifest elevated rates of joint hypermobility (50%) compared to the general population rate of 20% and a matched control population of 10%. Odds ratio for hypermobility in individuals with neurodevelopmental diagnoses, compared to the general population was 4.51 (95%CI 2.17-9.37), with greater odds in females rather than males. Neurodevelopmental patients reported significantly more symptoms of orthostatic intolerance and musculoskeletal skeletal pain than controls

**Conclusions:** In adults with neurodevelopmental conditions, there is a strong link between the expression of joint hypermobility, autonomic dysfunction and pain, more so than in healthy controls. Increased awareness and understanding of this association may enhance the management of core symptoms and allied difficulties including comorbid stress-sensitive physical symptoms.

## Introduction

Autism refers to a set of neurodevelopmental conditions (autism spectrum conditions, ASC) typically affecting social and emotional interaction, perception (with hypo- and hypersensitivities) and behaviour (often fixative obsessionality with change intolerance). Overlap with other neurodevelopmental conditions is common including up to 2/3^rd^ co-morbidity with attention deficit hyperactivity disorder (Davis and Kollins, 2012) and developmental tic disorder (e.g. Tourette Syndrome, in which there is 60% overlap with other neurodevelopmental conditions (Huisman-van Dijk et al., 2016)). In addition to the defining psychological features, autism and related neurodevelopmental conditions are recognised to carry increased vulnerability to physical health problems. These include stress sensitive disorders such as fibromyalgia, irritable bowel syndrome and fatigue and well as less clearly defined symptoms related to autonomic dysregulation, such as orthostatic intolerance. An association between autism and joint hypermobility is recognised that might elucidate vulnerability to physical symptoms (Baeza-Velasco et al. (2018b).

Joint hypermobility describes the ability of joints to move beyond typically ‘normal’ limits, usually as a consequence of ligamentous laxity, as occurs in connective tissue disorders or other genetic conditions (Clinch et al., 2011). In the general population, joint laxity is relatively common, yet prevalence can be difficult to estimate. One report suggests approximately 10% of the United Kingdom population have joint hypermobility (Sobey, 2015); another showed 28% of girls and 11% of boys (among 6,022 children) had generalised joint laxity (Clinch et al. (2011).

When joint hypermobility is associated with other symptoms, notably pain or autonomic dysfunction (Sobey, 2015, Clinch et al., 2011) a diagnosis of hypermobility spectrum disorder (HSD, formerly known as joint hypermobility syndrome) or Ehlers-Danlos Syndrome (EDS) may be made. Thirteen types of EDS have been described, with hypermobile EDS [hEDS--previously known as EDS-III], the most common (Demmler et al., 2019; Sobey, 2015).

Clinical features of HSD and hEDS are not limited to musculoskeletal and cutaneous symptoms (Demmler et al., 2019). Associations exist with cardiovascular autonomic dysfunction (Hakim et al., 2017b), gastrointestinal difficulties (Fikree et al., 2017), fatigue (Hakim et al., 2017a) and pain syndromes (Chopra et al., 2017), gynaecological and obstetric problems (Hugon-Rodin et al., 2016, Pezaro et al., 2018) and mental health concerns (Bulbena et al., 2017, Cederlöf et al., 2016).

As noted, in addition to the aforementioned conditions, there is growing recognition that hypermobility is associated with an increased likelihood of having comorbid neurodevelopmental conditions, including autism and attention deficit hyperactivity disorder (ADHD) (Bulbena et al., 2017, Cederlöf et al., 2016). Individuals with EDS are reported to be 7.4 times (95% CI: 5.2-10.7) more likely to have autism than controls (Cederlöf et al. (2016). Children with autism were shown to have greater mobility of joints (maximum passive joint mobility in degrees of angle were measured for a finger, wrist, elbow and ankle) compared to matched neurotypical controls (Shetreat-Klein et al. (2014) and the association between autism joint hypermobility syndrome/hEDS is further illustrated in a series of case studies (Sinibaldi et al., 2015), highlighting the need for more systematic research for robust characterisation of these links (Bulbena et al., 2017). ADHD is also associated with generalised joint hypermobility: One study reported generalised hypermobility in 32% of 54 ADHD patients, compared to 14% of controls. (Doğan et al. (2011). Another study reported the prevalence of generalised joint hypermobility to be 74% of 86 children with ADHD, compared to 13% of neurotypical controls (Shiari et al. (2013). Moreover, in a population-based matched cohort study in Sweden (n = 1,771), individuals with EDS were 5.6 times (95% CI: 4.2-7.4) more likely to have ADHD than those without EDS (Cederlöf et al. (2016).

Both autism and ADHD are commonly associated with the expression of tics. There has been so far no specific evidence published that links primary developmental tic disorder, exemplified by Tourette Syndrome, to joint hypermobility. Nevertheless, the clinical overlap and co-morbidity across neurodevelopmental conditions, suggests that people with Tourette Syndrome are more likely to manifest joint hypermobility compared to the general population. For example, there are high comorbidity rates of both ADHD and autism with TS, with estimates ranging from 60-80% for ADHD and 6.5-50% for autism (Cavanna et al., 2009) Interestingly, this neurodevelopmental triad also share a common involvement of fronto-striatal systems in respective symptomology(Rapanelli et al., 2017, Israelashvili et al., 2020) Comorbidity aside, even those with a single diagnosis of TS may be more predisposed to joint hypermobility than the general population given the links between autonomic dysfunction, premonitory urge sensations, and tics (Rae et al., 2019, Hawksley et al., 2015)

Orthostatic intolerance is a particular common expression of cardiovascular autonomic dysfunction in people with hypermobility conditions; symptoms such as light-headedness occur upon standing upright, and can be relieved by recumbence (Stewart, 2013). Symptoms of orthostatic intolerance can be debilitating (Hakim et al., 2017b). Associated syncope, fatigue, and migraines add to clinical burden and reduce quality of life (De Wandele et al., 2014).

Orthostatic intolerance is significantly more prevalent in individuals with joint hypermobility or hEDS compared to healthy controls and linked to postural tachycardia syndrome (PoTS) (Roma et al. (2018). PoTS is characterized by accelerating heart rate on standing and is one of the most common manifestations of orthostatic intolerance (Celletti et al. (2017). Orthostatic intolerance is observed in 80% of 35 JHS/EDS-III patients during autonomic testing, half of whom met criteria for postural orthostatic tachycardia syndrome. Furthermore, in a survey of 116 patients diagnosed with JHS/EDS-III, nearly everyone (98%) reported the experience of orthostatic intolerance, experiencing dizziness when getting out of bed in the morning or when exercising in the heat or during/after a hot shower (Chan et al., 2019). Therefore, orthostatic intolerance is frequent in people with hypermobility.

Given the co-occurrence of hypermobility and neurodevelopmental conditions, it is reasonable to hypothesise that individuals with neurodevelopmental conditions are more likely to experience orthostatic intolerance compared to controls. Moreover, a more direct association is proposed between orthostatic hypotension and specific behavioural and emotional disorders (Perlmuter et al. (2013). Correspondingly, in healthy preschool children, poorer pulse pressure regulation during orthostatic challenge (a measure of orthostatic hypotension) predicts higher ADHD scores (Casavant et al. (2012). Moreover, children with poorer responses to orthostatic stress were more likely to be viewed as oppositional or have attention difficulties (Casavant et al. (2012). While this study did not test ADHD patients, it points towards a relationship between orthostatic intolerance and ADHD which warrants further scrutiny.

Similarly, minimal systematic research in this domain has involved individuals with autism or Tourette Syndrome: One case series found 5 of six patients with autism (aged 12 to 28) had significant orthostatic intolerance on autonomic function testing. (Goodman (2016). Conversely, no clinical signs of dysautonomia during orthostatic challenge (head up tilt) test were observed in 39 boys with autism (Bricout et al., 2018). Case studies provide evidence of an association between orthostatic hypotension and Tourette Syndrome (Minns et al., 2010). Yet clearly more research involving larger samples is required to characterise fully relationships across neurodevelopmental disorders (including autism and Tourette Syndrome) with orthostatic intolerance and dysautonomia.

Pain is a common symptom in individuals with hypermobility (Chopra et al., 2017). Given the observed association between neurodevelopmental conditions and hypermobility, individuals with autism, ADHD, and Tourette Syndrome may have an increased likelihood of experiencing pain (Baeza-Velasco et al., 2018a). For instance, an online survey of autistic women found that 100% of those with joint hypermobility (n = 85) experienced joint pain, compared to only 29% of those without joint hypermobility (n = 20). Chronic pain has a detrimental effect on quality of life in individuals with neurodevelopmental conditions (Asztély et al., 2019) highlighting a need to further characterise these associations within a more representative sample including males (Baeza-Velasco et al., 2018a). In ADHD, where the link to hypermobility related disorders is arguably more established (Baeza-Velasco et al., 2018b), the experience of pain may interact with, and impact negatively, attention (Moore et al., 2019). In a study of referrals to a pain clinic, ADHD patients reported statistically significantly higher mean pain scores compared to non-ADHD patients (Kasahara et al. (2020). While over three-quarters of women with ADHD and/or autism report chronic pain (Asztély et al., 2019) this study did not test if hypermobility was a mediating factor.

### Aims and hypotheses

Motivated by the evidence described above of associations between hypermobility, orthostatic intolerance and pain symptoms in individuals with neurodevelopmental conditions, this study tested whether group differences exist between individuals with diagnoses of neurodevelopmental conditions (patients with autism, ADHD and Tourette Syndrome) and neurotypical controls on measures of generalised joint laxity, autonomic symptoms, and pain. We hypothesized that, compared to neurotypical individuals, a significantly greater proportion of neurodevelopmental patients will have generalised joint laxity, and experience significantly more symptoms of orthostatic intolerance and musculoskeletal pain. We further hypothesised that more women compared to men would express generalised joint laxity, as is commonly found, and that greater generalised joint laxity would predict increased orthostatic intolerance and pain.

## Method

### Participants

Data was analysed from 138 participants in total. One hundred and nine patient participants with neurodevelopmental conditions were assessed in total. Eighty-seven were assessed as part of a study of prevalence of hypermobility and autonomic symptoms in secondary care outpatient clinics in Sussex, including a specialist Neurodevelopmental Service (n=87, NRES Ethics Committee (South East Coast) (12.LO.1942)). For these patient participants recruited directly from secondary care mental health clinics, the clinical notes of patients were evaluated to confirm that they had received a clinical diagnosis of autism, ADHD, or Tourette Syndrome. Additional individuals with Tourette syndrome (n=22) were recruited from a separate study ((NRES Ethics Committee (South East Coast) (15.LO.0109)). See Table 2 for individual rates of disorder and co-morbidity. These neurodevelopmental patient participants were compared to healthy controls (n=29), recruited to a study to validate the autonomic symptoms questionnaire measure (An autonomic and quality of life self-administered questionnaire (AQQoL) (Iodice, 2013)) Participants in this control group were adults with no diagnosed neurodevelopmental, mental health or neurological conditions.

Eligibility criteria for adult patient participants included confirmed diagnosis of neurodevelopmental condition (autism, ADHD, Tourette Syndrome).

### Measures and materials

The autonomic symptoms questionnaire measure (an autonomic and quality of life self-administered questionnaire (AQQoL)) (Iodice, 2013) was used to measure symptoms of orthostatic intolerance and musculoskeletal symptoms, and included the clinician-rated Beighton scoring system. The 9-item Beighton Scoring System (Beighton et al., 1973) was used to assess participants’ joint laxity. Scores of 4 and above were indicative of generalised joint laxity.

Participants were asked to rate on a 5-point Likert scale (from ‘no’ to ‘yes -daily’) how often they experienced various symptoms of orthostatic intolerance (e.g. ‘do you feel dizzy or lightheaded?’). Incidence of musculoskeletal symptoms (predominately pain) were also surveyed (e.g. ‘do you have any of the following symptoms? a) pains in the knees b) pains in the fingers’) Participants were asked to rate these questions on a 3-point Likert scale in relation to frequency (from ‘no’ to ‘yes - for longer than 3 months’). All participants completed this full questionnaire, except the patient participants in the Tourette Syndrome study who only completed the Beighton Scoring and orthostatic intolerance subscale of the AQQoL.

### Data Preparation for Statistical Analyses

In the original validation of the AQQoL, the Beighton score was included in the total musculoskeletal subscale. Here, the Beighton score was analysed separately as it was hypothesised that there would be a difference in Beighton scores between patients and controls.

Chi-square tests were used to test for group differences between neurodevelopmental patients and controls, in the proportion of people with generalised joint laxity. Expected frequencies were tested to ensure the use of Pearson’s chi-square was appropriate.

Mann-Whitney U tests were used to test for group differences between neurodevelopmental patients and controls, and compare: 1) Beighton scores, 2) orthostatic intolerance symptom scores, and 3) musculoskeletal scores. This approach was used because scores on these three measures for each group were not normally distributed. Binary logistic regression was used to calculate the odds ratios of having generalised joint laxity for patients compared to the general population. Spearman’s correlations were also calculated between Beighton scores and: 1) orthostatic intolerance symptom scores and 2) musculoskeletal scores given the predicted relationships between these (i.e. higher Beighton scores will be positively correlated with increased orthostatic intolerance symptom scores and musculoskeletal scores). Binary logistic regression was used to calculate odds ratios in relation to the likelihood of having generalised joint laxity across genders and to compare rates of joint laxity in our sample to the general population (Clinch et al., 2011)

## Results

Overall, 82 participants were male (59%) and the average age was 35.5 years old (SD = 13.0). Sixty-seven of patients were male (62% of this group) and fifteen of the control group were male (52% of this group). The average age of patients was 34.9 years old (SD = 11.3) and the average age of controls was 37.9 years old (SD = 17.9). There were no significant differences between patients and controls in terms of age or sex (Table 1).

**Table 1.** Demographic variables of patients and controls.

| Characteristics | Patients (n = 109) | Controls (n = 29) | Difference in demographics between groups |
| --- | --- | --- | --- |
| Age in years (M, SD) | 34.9 (11.3)<br>Range: 18-61 | 37.9 (17.9)<br>Range: 19-68 | p = .746 |
| Sex, male (% of group) | 67 (62%) | 15 (52%) | p = .342 |
| Beighton score /9 (M, SD) | 3.2 (2.6) | 0.9 (2.0) | p=<0.001 |
| Orthostatic intolerance symptom score (M, SD) | 24.2 (15.6) | 4 (4.3) | p=<0.001 |
| Musculoskeletal score | 6.8 (3.7) | 2.8 (2.7) | p=<0.001 |

### Neurodevelopmental and mental health diagnoses

Thirty patients had a diagnosis of autism (28% of patients). sixty-seven of the patients were diagnosed with ADHD (62% of patients), and twenty-four patients had Tourette Syndrome (22% of patients). Twenty-seven patients had comorbid diagnoses of autism and ADHD, (25% of patients) nine patients were diagnosed with ADHD and Tourette Syndrome (9% of patients).

**Table 2.**
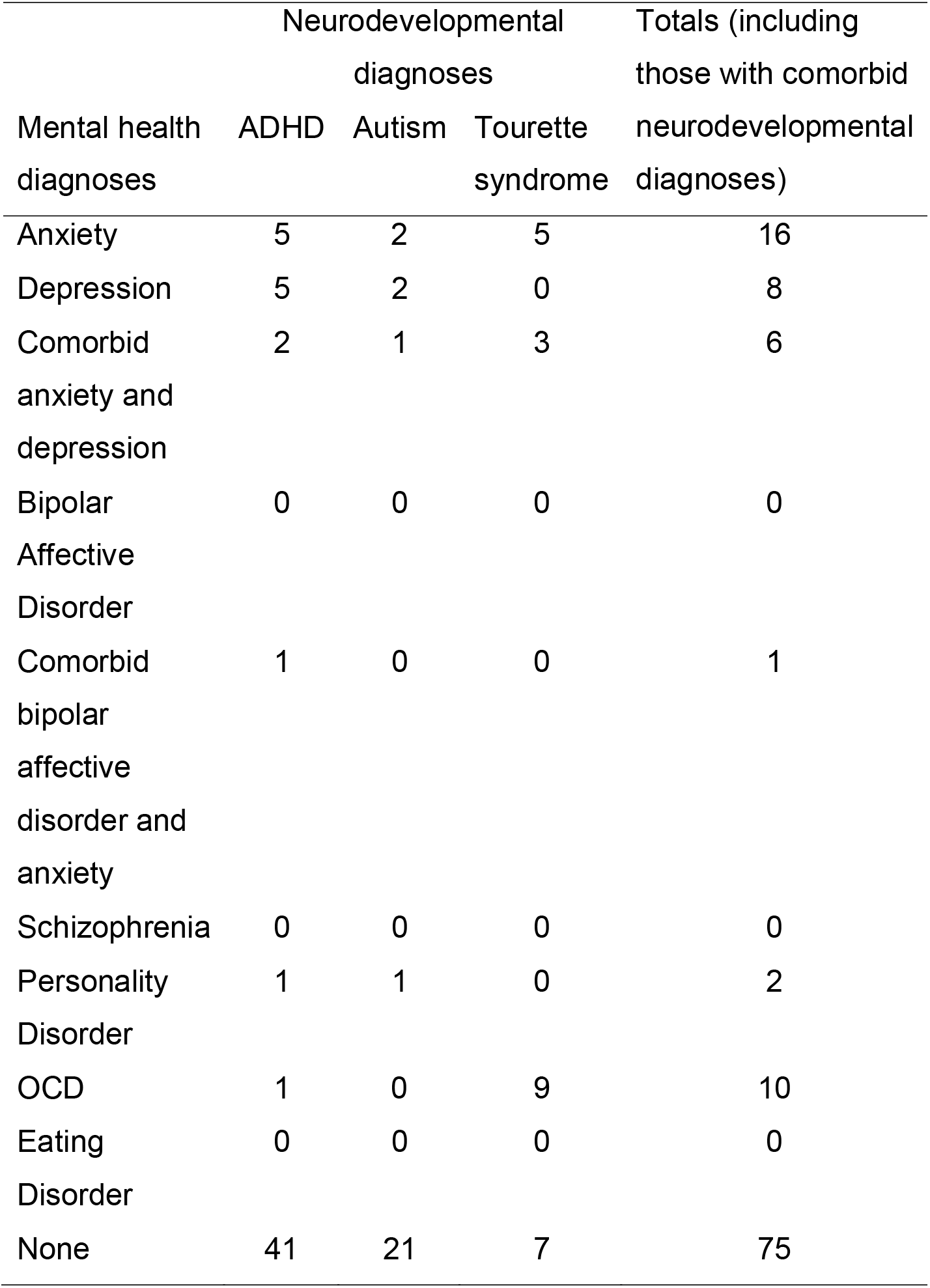
Diagnosed neurodevelopmental and mental health conditions in patients

### Generalised joint laxity

Fifty-four (50%) of the patient group met criteria for generalised joint laxity compared to 3 (10%) of the control group (Table 3). See Figure 1 for percentage breakdown per condition in terms of hypermobility. Data from the ALSPAC birth cohort (Clinch et al., 2011), was used to compute the odds ratio of having generalised joint laxity if individuals had an diagnosis of a neurodevelopmental condition, compared to the general population: Generalised joint laxity was 4.51 (95% CI: 2.17, 9.37) times higher if individuals were autistic, 4.34 (95% CI: 2.67-7.03) times higher if individuals had ADHD and 7.02 (95% CI: 3.06-16.1) times higher if individuals had Tourette Syndrome.

**Figure 1.**
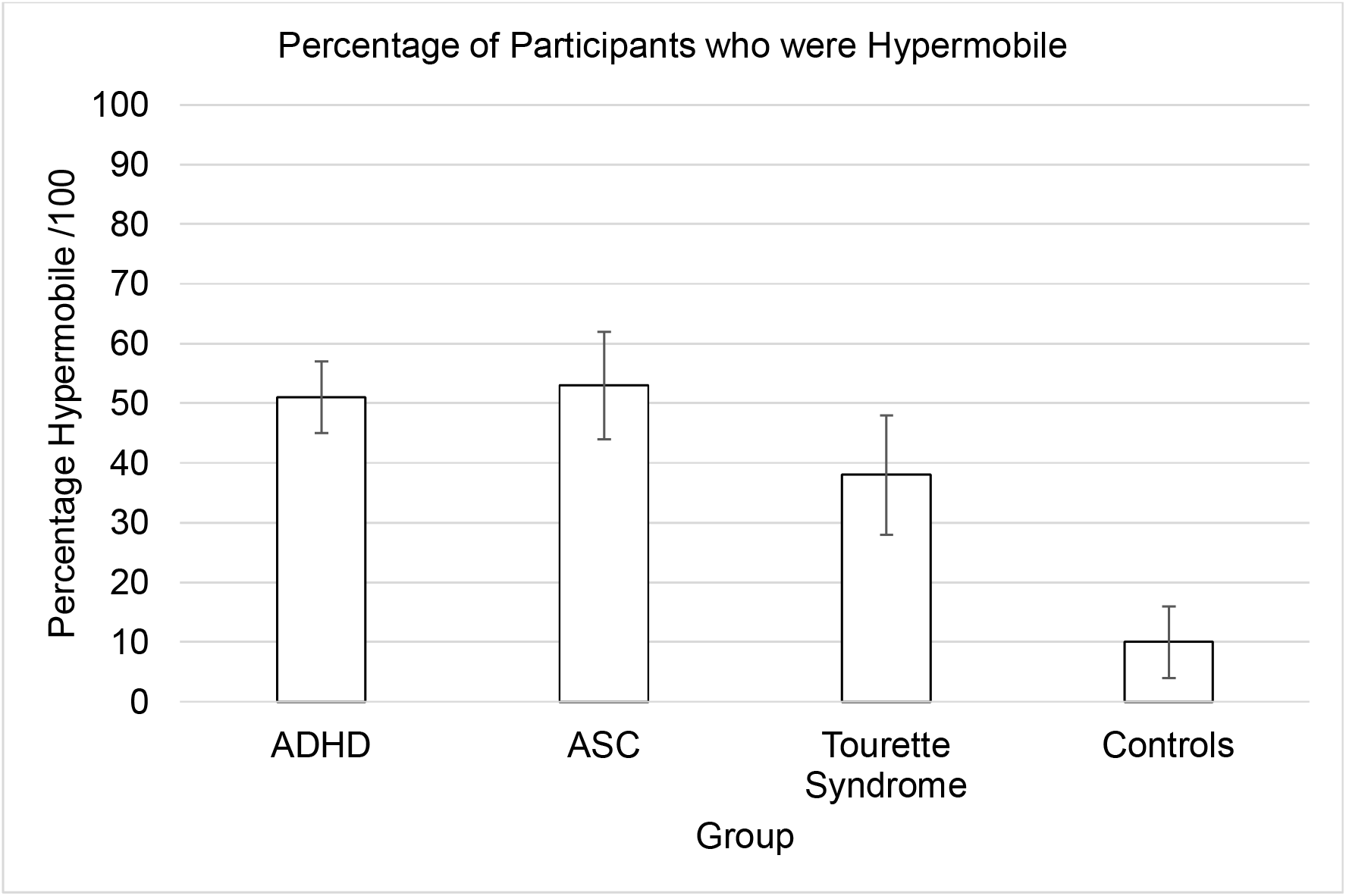
Percentage of individuals in each group who had generalised joint laxity. Error bars show 95% CI. Beighton score was significantly higher *(Mdn* = 4) in patients compared to controls (Mdn = 0), *U* = 2459, *z* = 4.69, *r* = 0.40 *p=* <0 .001, (see Table 1 for mean scores).

In neurodevelopmental patients in our cohort, there was also a significant association between gender and whether they had generalised joint laxity or not, that is females were more likely to have generalised joint laxity x^2^(1) = 9.63, *p* = .002 (see figure 2). Based on the odds ratio, the odds of patients having generalised joint laxity were 3.52 (95% CI: 1.55, 7.97) times higher if they were female compared to male.

In all participants, including our controls, there was a significant association between gender and whether individuals had generalised joint laxity or not: Females were more likely to have generalised joint laxity x^2^(1) = 6.88, *p* = .009. The odds of a participant having generalised joint laxity was 2.53 (95% CI: 1.26, 5.09) times higher if they were female compared to male.

**Figure 2.**
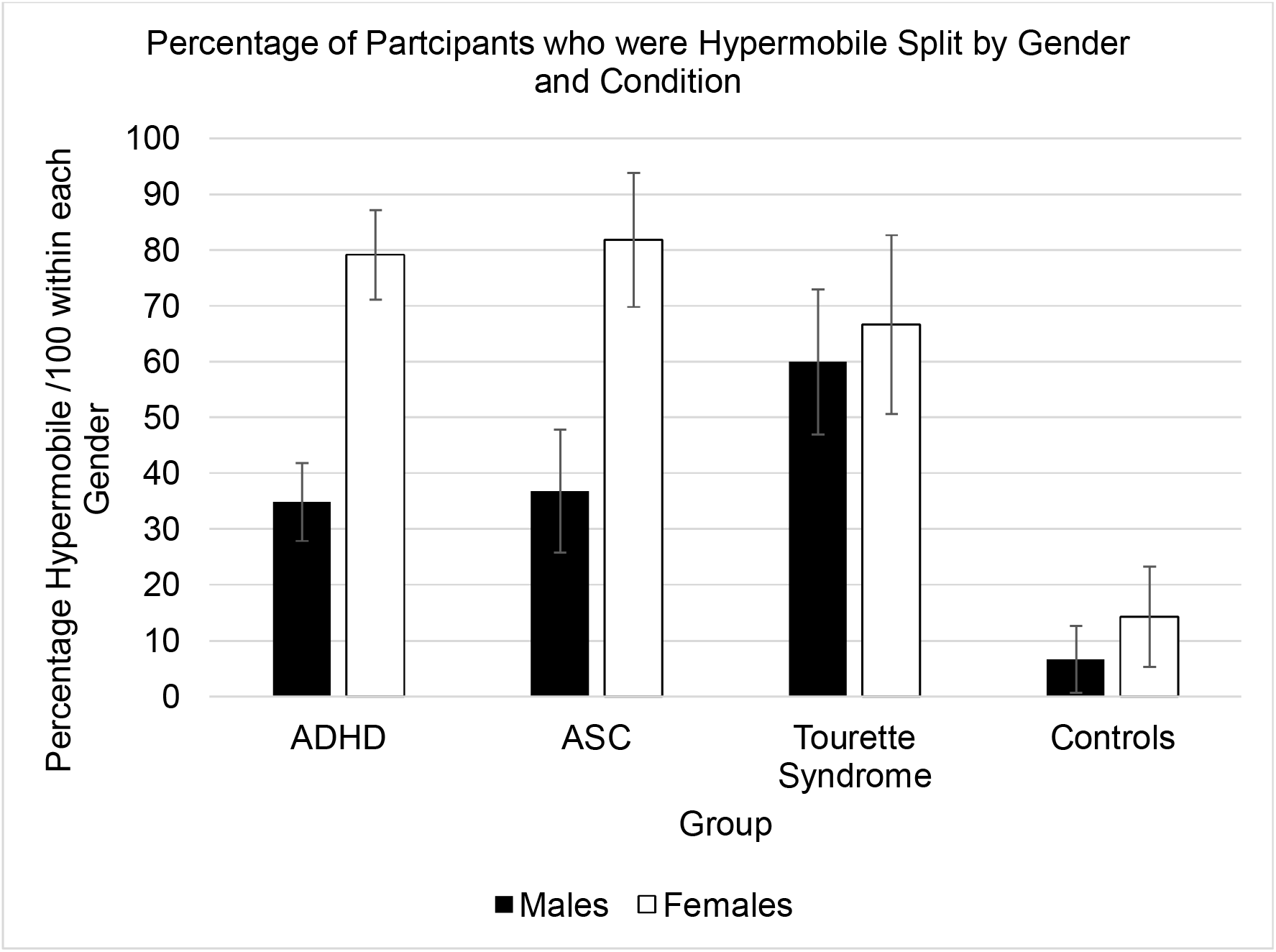
Percentage of participants who had generalised joint laxity within each gender across conditions. Error bars show 95% CI.

### Orthostatic intolerance

Orthostatic intolerance symptom score in patients was significantly higher *(Mdn* = 21.5) compared to controls *(Mdn* = 3), *U* = 2829.5, *z* = 7.21, *r* = 0.63, *p* < 0.001, (Figure 3).

**Figure 3.**
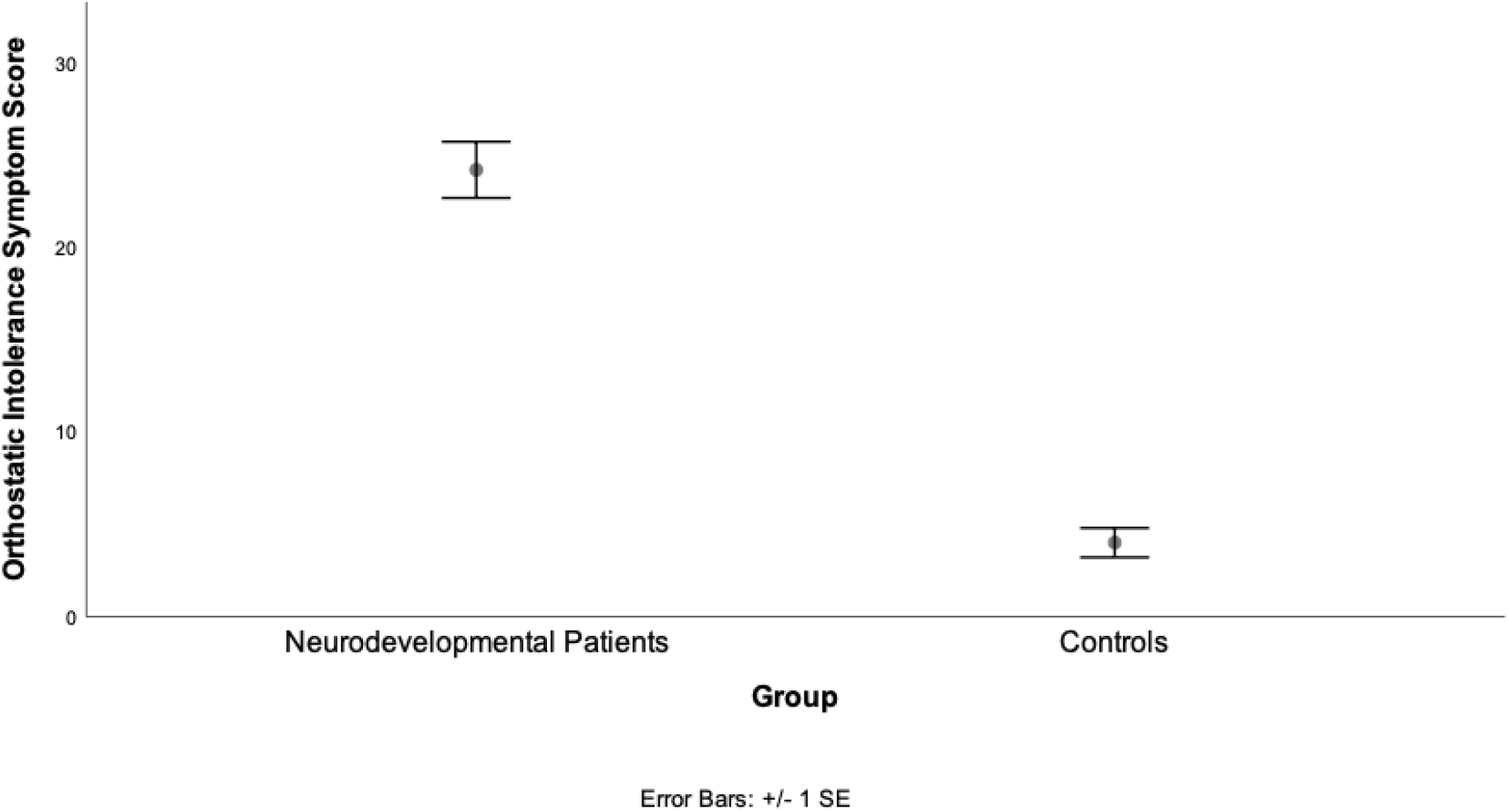
Difference in mean orthostatic intolerance symptom score between neurodevelopmental patients and controls

We observed a positive correlation between orthostatic intolerance symptom score and generalised joint laxity (p = < 0.001). This suggests that the higher the participants’ Beighton score is, the greater their experience of symptoms of orthostatic intolerance (Figure 4).

**Figure 4.**
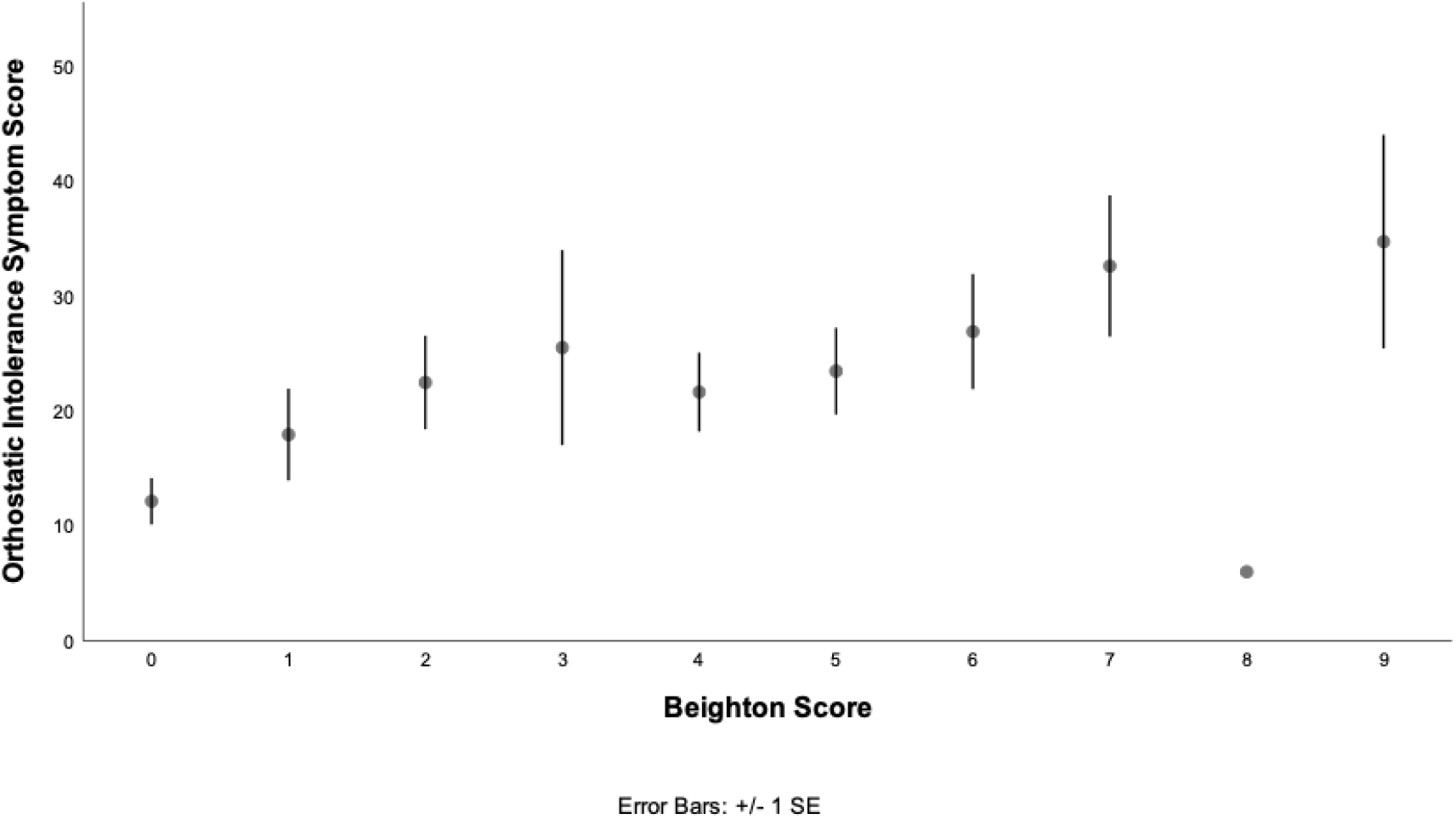
Graph showing the relationship between orthostatic intolerance symptom score and Beighton score

### Musculoskeletal symptoms

Musculoskeletal symptom score was significantly higher in patients *(Mdn* = 6) compared to controls *(Mdn* = 3), U = 1811.5, *z* = 4.95, *r* = 0.48 *p* < 0.001, (Figure 5).

**Figure 5.**
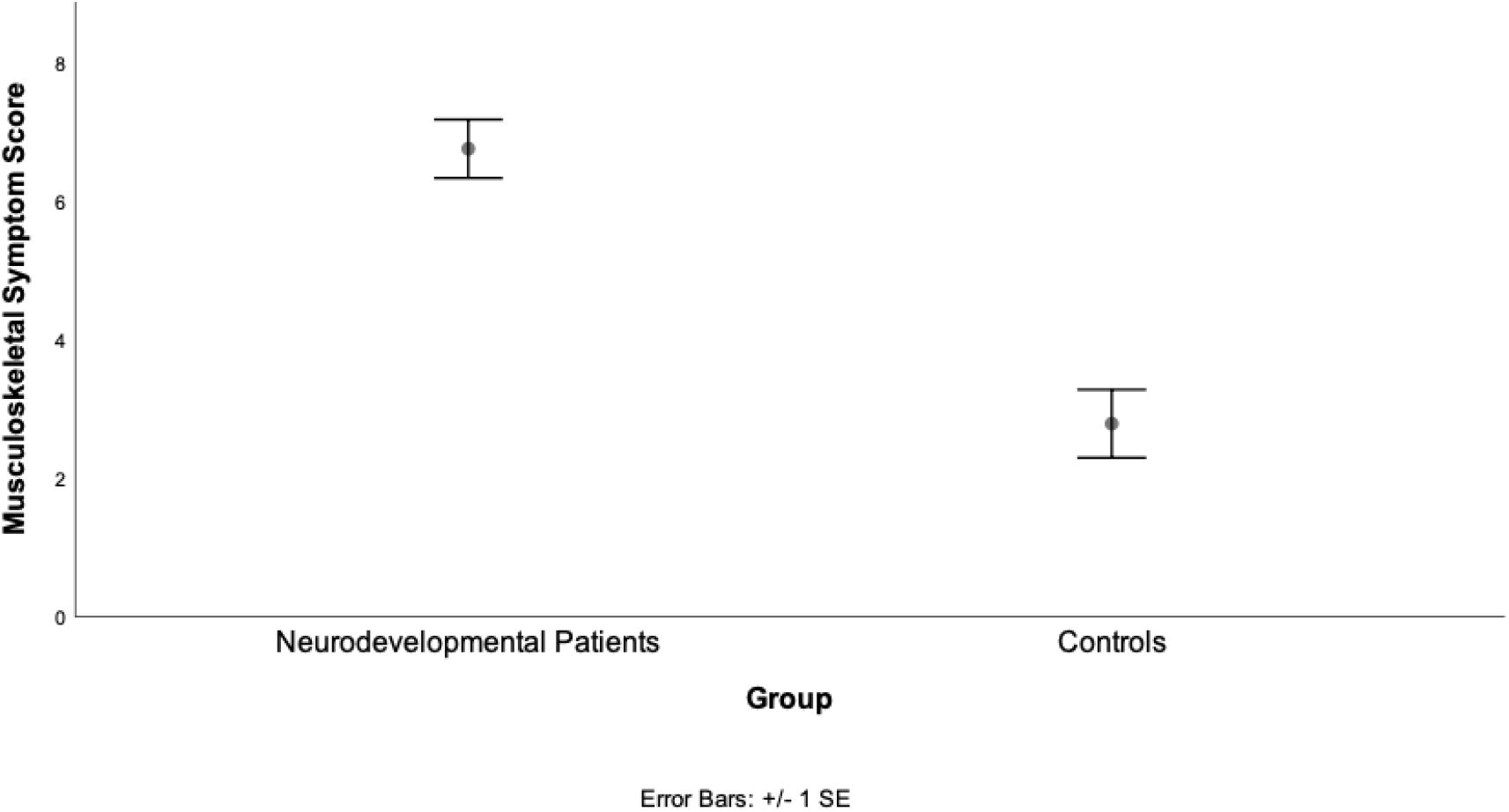
Difference in mean musculoskeletal symptom scores between neurodevelopmental patients and controls

A positive correlation was observed between musculoskeletal symptom score and hypermobility (*p* = .001). This suggests that the greater the participants’ Beighton score, the higher their musculoskeletal symptom score. (Figure 6).

**Figure 6.**
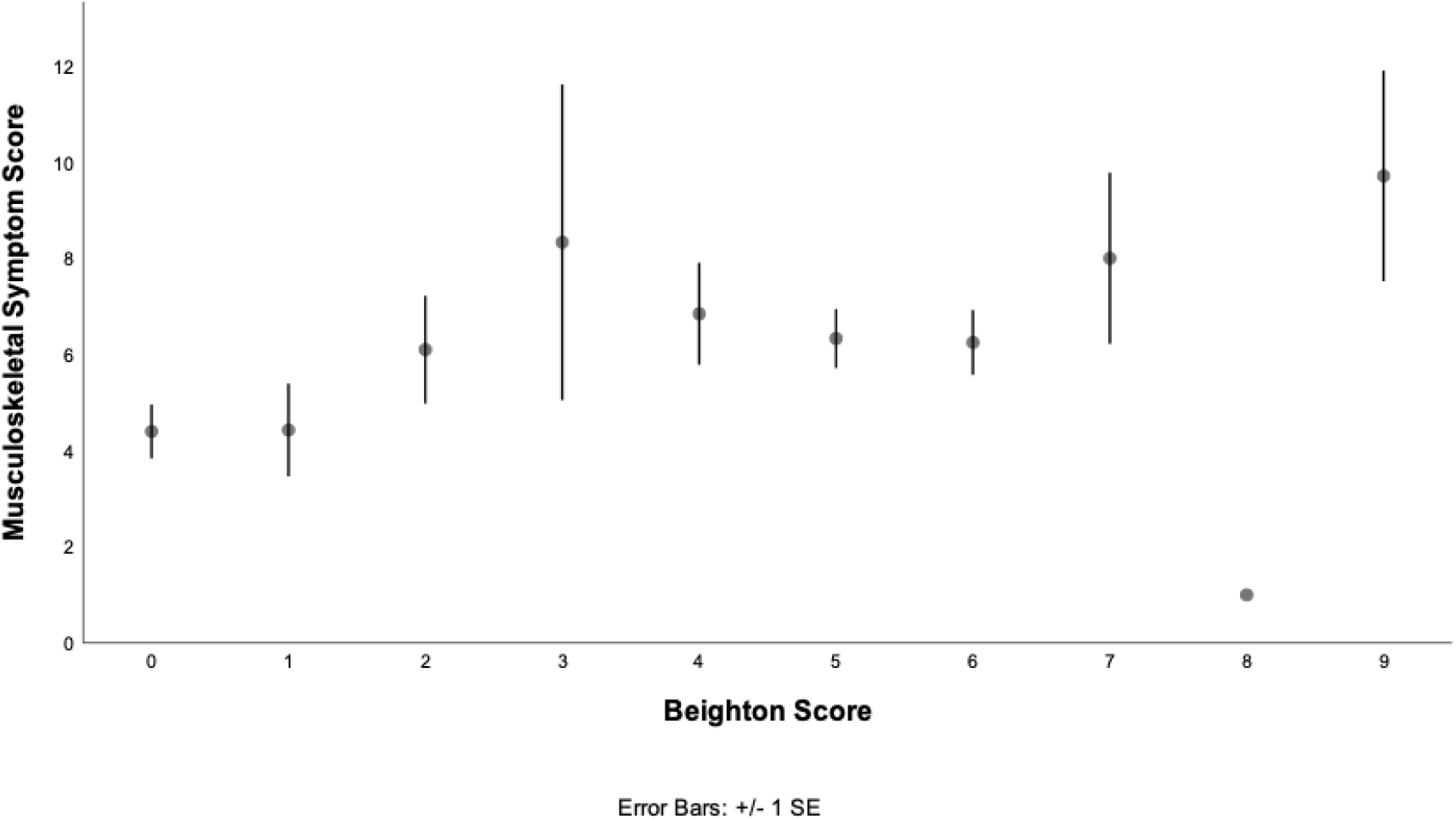
Graph showing the relationship between musculoskeletal symptom score and Beighton score

## Discussion

This is the first study to investigate the prevalence of generalised joint laxity, orthostatic intolerance symptoms and pain (in the form of musculoskeletal symptoms) across adult neurodevelopmental patients (including Tourette Syndrome) compared to healthy controls. All hypotheses were supported, such that neurodevelopmental patients were significantly more likely to have generalised joint laxity, experience orthostatic intolerance and pain. Generalised joint laxity was more common in women and was associated with both orthostatic intolerance and pain.

Half of patients with a neurodevelopmental condition had generalised joint laxity compared to just 10% of controls, extending brain imaging findings in hypermobility that link to possible neurodevelopmental conditions (Eccles et al., 2012). The finding that 53% of these adult patients with autism had generalised joint laxity extends a growing literature from case studies, children and cohort research (Sinibaldi et al., 2015, Shetreat-Klein et al., 2014, Cederlöf et al., 2016). Fifty-one percent of ADHD patients had generalised joint laxity which falls between the lower estimate in previous research with adults (32% of 54 patients (Doğan et al., 2011)) and the higher estimate in research with children (74% of 86 patients (Shiari et al., 2013). Demonstrating 38% of Tourette Syndrome patients had generalised joint laxity provides the first reported systematic estimate of this association. This observation highlights the increased prevalence of generalised joint laxity across neurodevelopmental conditions.

The significantly higher prevalence of orthostatic intolerance symptoms of neurodevelopmental patients compared to controls provides direct empirical evidence for this suggestion within the literature (Perlmuter et al., 2013, Goodman, 2016). Moreover, our larger study used quantitative data from patients with diagnosed neurodevelopmental conditions, extending an association between orthostatic symptoms and ADHD questionnaire score in fewer healthy individuals (Casavant et al., 2012) as opposed to confirmed diagnosis. We observed increased orthostatic intolerance across patients with autism, ADHD, and Tourette Syndrome. The relationship with hypermobility may partly explain why orthostatic intolerance is more prevalent in patients with neurodevelopmental conditions given the significant association observed here between orthostatic intolerance symptom score and Beighton score.

Higher pain symptom experience in neurodevelopmental patients compared to controls has also previously been suggested in the literature (e.g. Kasahara et al. (2020). We confirmed a similar average amount of musculoskeletal symptoms reported by patients across neurodevelopmental conditions as earlier studies. Compared to past studies, the data from our study encompassed more men, which enables conclusions to more generalisable since autism and ADHD are more commonly diagnosed in men. It is likely that hypermobility leads to increased likelihood of pain given the correlations between Beighton score and musculoskeletal symptoms, and the high prevalence of hypermobility reported in this sample.

This study provides compelling evidence for increased rates of joint hypermobility - a constitutional variant in systemic connective tissue in individuals with psychiatric diagnoses of neurodevelopmental condition. Orthostatic intolerance - an expression of autonomic cardiovascular dysregulation and musculoskeletal pain were also shown to be expressed at higher rates across individuals with neurodevelopmental conditions and showed a clear relationship to joint laxity. Differences in connective tissue may be a mediating factor for both these associations, causing pooling of blood in lax peripheral vessels (as in PoTS(Mathias et al., 2011)) and enhancing sensitivity to tissue stretch and damage. Here, we highlight the relevance in the context of a set of neurodevelopmental conditions in where differences in perceptual sensitivity, abnormalities in proprioception and interoception(Rae et al., 2019, Garfinkel et al., 2016) contribute to both the psychosocial expression of these conditions but also to the (often under recognised) vulnerabilities to chronic physical symptoms which further decrease quality of life. Autism ADHD and developmental tic disorder (Tourette syndrome) often share often complex symptoms, consistent with overlapping neurodevelopmental aetiopathology. Increasingly, services need to recognise such complexity and move beyond ‘exclusive’ diagnostic categories and traditional boundaries between physical and mental health (Eccles et al., 2020) This paper sets the scene for further studies that test explicitly for meditating interactions to enable targeting of interventions to enhance quality of life across psychological and physical domains.

## Data Availability

Deidentified data is available upon reasonable request from

